# The Role of HIV Partner Services in the Modern Biomedical HIV Prevention Era: A Network Modeling Study

**DOI:** 10.1101/2022.05.20.22275395

**Authors:** Samuel M. Jenness, Adrien Le Guillou, Cynthia Lyles, Kyle T. Bernstein, Kathryn Krupinsky, Eva A. Enns, Patrick S. Sullivan, Kevin P. Delaney

## Abstract

**Background:** HIV partner services can accelerate the use of antiretroviral-based HIV prevention tools (ART and PrEP), but its population impact on long-term HIV incidence reduction is challenging to quantify with traditional PS metrics of partner identified or HIV-screened. Understanding the role of partner services within the portfolio of HIV prevention interventions, including using it to efficiently deliver antiretrovirals, is needed to achieve HIV prevention targets.

**Methods:** We used a stochastic network model of HIV/STI transmission for men who have sex with men (MSM), calibrated to surveillance-based estimates in the Atlanta area, a jurisdiction with high HIV burden and suboptimal partner services uptake. Model scenarios varied successful delivery of partner services cascade steps (newly diagnosed “index” patient and partner identification, partner HIV screening, and linkage or reengagement of partners in PrEP or ART care) individually and jointly.

**Results:** At current levels observed in Atlanta, removal of HIV partner services had minimal impact on 10-year cumulative HIV incidence, as did improving a single partner services step while holding the others constant. These changes did not sufficiently impact overall PrEP or ART coverage to reduce HIV transmission. If all index patients and partners were identified, maximizing partner HIV screening, partner PrEP provision, partner ART linkage, and partner ART reengagement would avert 6%, 11%, 5%, and 18% of infections, respectively. Realistic improvements in partner identification and service delivery were estimated to avert 2–8% of infections, depending on the combination of improvements.

**Conclusions:** Achieving optimal HIV prevention with partner services depends on pairing improvements in index patient and partner identification with maximal delivery of HIV screening, ART, and PrEP to partners if indicated. Improving the identification steps without improvement to antiretroviral service delivery steps, or vice versa, is projected to result in negligible population HIV prevention benefit.

## INTRODUCTION

HIV partner services has the potential to prevent HIV by intervening with the exposed partners of persons newly diagnosed with HIV (index patients). Recommended partner interventions include HIV screening to diagnose asymptomatic infection, with subsequent linkage to HIV services depending on the test result.^1^ Traditionally in the United States, HIV partner services activities have focused on linkage to antiretroviral therapy (ART) for partners who are newly diagnosed,^2^ potentially lowering community HIV viral load (VL) and reducing rates of subsequent HIV transmission (i.e., “treatment as prevention”).^3^ However, some jurisdictions have used HIV partner services as a pathway to other prevention services, such as HIV preexposure prophylaxis (PrEP) and STI screening.^4^

HIV partner services consistently increase the coverage of these HIV services,^5^ but the degree to which this program can contribute to population-level HIV incidence reduction is unclear. Achieving this population impact requires reducing the time to ART or PrEP uptake compared to what is achieved through other, individual-level interventions. For example, the average time from HIV infection to VL suppression is 44 months.^6^ For partner services to reduce HIV transmission rates, it must identify a large fraction of HIV-infected partners currently unsuppressed and link them to ART within this period.^7,8^ Current partner services metrics suggest challenges in achieving this.^1^

Gay, bisexual and other men who have sex with men (MSM) have been the largest group of users of HIV partner services in the U.S. due to their disproportionate HIV incidence.^1^ While levels of ART and PrEP coverage have increased among MSM, gaps between actual and recommended use persist, particularly in disadvantaged MSM subpopulations.^9^ Optimization of partner services for these MSM may be an approach to improving the uptake of these biomedical tools, particularly in jurisdictions (such as Atlanta) where the current partner services infrastructure is weak and the incidence of HIV is high.^10,11^

In this study, we used a stochastic network model of HIV transmission dynamics among MSM in Atlanta to evaluate the role of HIV partner services in this current HIV biomedical prevention era. Our aims were to quantify how both traditional (HIV screening and ART linkage) and newer (PrEP linkage) components of HIV partner services function together to achieve HIV prevention goals.

## METHODS

### Study Design

We used a stochastic network model of HIV, *Neisseria gonorrhoeae* (NG), and *Chlamydia trachomatis* (CT) transmission dynamics for U.S. MSM, built with the *EpiModel* software platform.^12^ Our model simulates disease dynamics over dynamic sexual partnership networks using temporal exponential random graph models (TERGMs).^13^ This approach is necessary to realistically represent and track sexual partnership histories. This current study builds on our previous studies of partner-based strategies for HIV/STIs.^14^ Full methodological details are provided in an Appendix [**LINK**], and model code is available on Github [http://github.com/EpiModel/CombPrevNet].

Our model represented main, casual, and one-time sexual partnerships for Black, Hispanic or Latino (Hispanic), and White/Other-race (White) MSM, aged 15 to 65, in the Atlanta metropolitan statistical area. The starting network size in model simulations was 10,000 MSM (approximately one-tenth of the estimated Atlanta MSM population size of 102,000^15^), which could increase or decrease over time based on arrival (sexual debut) and departure (mortality or sexual cessation) (**Appendix S5**).

### Core HIV/STI Model

The model consisted of five core components: 1) statistical network models (TERGMs) to represent dynamic sexual partnership formation and dissolution; 2) statistical generalized linear models (GLM) to predict behavior within partnerships; 3) simulation of pathogen transmission across active partnerships; 4) simulation of HIV/STI disease progression, recovery, and natural history; and 5) simulation of HIV/STI prevention and clinical interventions (including partner services). NG and CT transmission were simulated along the same sexual network as HIV, but with disease recovery. Full details on the STI components are found in **Appendix S9–12**.

Empirical data from the ARTnet study were used to fit the statistical models for networks and behavior (components 1 and 2) (**Appendix S2**). ARTnet was a web-based egocentric network study conducted in 2017–2019 of MSM in the US.^16^ We weighted model parameters by census-based race/ethnicity and age distributions for Atlanta MSM to account for the ARTnet sampling design. Predictors in TERGMs for partnership formation included partnership type (main, casual, and one-time), demographic heterogeneity in network degree, and assortative mixing by demographics and sexual position (**Appendix S3**). Partnership dissolution was modeled for main and causal partnerships, with rates stratified by partnership type and age. GLMs were fit to predict the frequency of sexual acts per partnership per week and the probability of condom use per act as a function of race/ethnicity, age, HIV status, and partnership type and duration (**Appendix S4**).

Our base model included both individual-based and partner services-based (described below) pathways to HIV screening, treatment, and prevention services (**Appendix S7**). For the individual-based pathway, in each weekly time step, MSM could be screened for HIV based on rates calibrated to reproduce estimates from the Georgia Department of Public Health (GDPH) surveillance of the proportion of MSM with HIV who were diagnosed.^17^ MSM who screened positive entered the HIV care cascade, with opportunities to initiate antiretroviral therapy (ART), which reduced vial load (VL) and disease progression. MSM on ART could cycle off and on ART based on rates also calibrated to GDPH data.^18^ Lower VL due to sustained ART use was associated with a reduced probability of HIV transmission. Other factors modifying the HIV transmission probabilities included PrEP use, condom use, sexual position, circumcision, and current NG or CT infection (**Appendix S8**).

MSM who screened HIV-negative entered the HIV prevention cascade, consisting of initiation, adherence, and persistence in PrEP care for daily oral tenofovir/emtricitabine.^19^ MSM who tested HIVnegative and met indications for PrEP based on CDC guidelines were eligible to start.^20^ Indicated MSM then started PrEP based on an initiation probability generating a coverage level of 15%.^9^ Heterogeneous PrEP adherence was modeled, with 78.4% meeting a high-adherence level that resulted in a 99% reduction in HIV risk.^21^ PrEP discontinuation was based on secondary estimates of the proportion of MSM who were retained in PrEP care at 6 months (57%).^22^ PrEP care included quarterly HIV and STI screening.^20^ *HIV Partner Services*. **Table 1** summarizes the design elements and parameters for partner services in our model. The four included steps of the partner services cascade were: 1) identification of newly diagnosed MSM (“index patients”) through routine HIV screening, and their participation in partner services; 2) elicitation and identification of recent sexual partners of index patients; 3) provision of HIV screening to those partners; and 4) linkage of partners to HIV care or prevention services, depending on partner HIV status. Below, we call the first two steps “upstream” and the latter two steps “downstream.” The base (calibrated) model scenario represents partner services as currently delivered in Atlanta, whereas the counterfactual (experimental) scenarios modify metrics for each step.

**Table 1.** Partner Services Cascade Parameters and Sources

| Parameter | Description | Base Model Parameter |  | Sources |
| --- | --- | --- | --- | --- |
|  |  | Missing = No | Complete Case |  |
| Index Network Degree | Minimum number of cumulative partners in window before triggering partner services | 1 | 1 | Standard Operating Protocol<sup>1</sup> |
| Index Initiation | Probability that an index patient will be initially offered and accept partner services | 66.6% | 66.6% | 2018 CDC Partner Services Report<sup>2</sup> |
| Index Partnership Window |  |  |  |  |
| *Main Partners* | Number of weeks in the past that a partnership of each type qualifies for partner elicitation | 52 | 52 | Standard Operating Protocol<sup>1</sup> |
| *Casual Partners* |  | 52 | 52 |  |
| *One-Time Partners* |  | 52 | 52 |  |
| Partner Identification |  |  |  |  |
| *Partners:Index Ratio* | Number of partners identified per index patient | 0.432 | 0.432 | 2018 CDC Partner Services Report<sup>2</sup> |
| *Casual to Main Scalar* | Odds ratio for the probability identification of casual partners to that of main partners | 0.50 | 0.50 |  |
| *One-Time to Main Scalar* | Odds ratio for the probability identification of one-time partners to that of main partners | 0.25 | 0.25 |  |
| Partner HIV Screening | Probability of completing HIV screening for partners identified and reached | 39.4% | 84.0% | 2018 CDC Partner Services Report<sup>2</sup> |
| Partner PrEP Linkage | Probability of a partner who screened HIV- and with current indications starting PrEP | 0 | 0 | Assumed based on Standard Operating Protocol for ATL<sup>1</sup> |
| Partner Care Linkage | Probability of a partner who newly screened HIV+ and not currently in care will start ART | 38.7% | 96.0% | CDC Partner Services Report<sup>2</sup> |
| Partner ART Reengagement | Probability of a partner who previously screened HIV+ and previously in ART care will restart ART | 0 | 0 | Assumed based on Standard Operating Protocol for ATL<sup>1</sup> |

<sup>1</sup> Communication with CDC co-authors (KB and CL) familiar with standard partner services protocol. <sup>2</sup> Available at: https://www.cdc.gov/hiv/pdf/library/reports/cdc-hiv-partner-services-annual-report-2018.pdf

All sexually active index patients (minimum cumulative network degree = 1) within the partnership window (timeframe = past 52 weeks) were eligible for partner services. In the base scenario, 67% of index patients accepted partner services (**Table 1**). Partners of index patients were identified with probabilities stratified by partnership type, highest for main partners and lowest for one-time partners. Identified partners could self-report as either HIV-positive or unknown status; the latter were offered HIV screening. Partners who newly screened HIV-positive or self-reported HIV-positive and were still ART-naïve were eligible for HIV care linkage, resulting in immediate ART initiation. Partners who screened HIV-negative were eligible for immediate PrEP linkage if they met CDC indications.^20^ Partners who selfreported as HIV-positive but who had fallen out of care were eligible for immediate ART reengagement. Our base scenarios assumed no PrEP linkage or ART reengagement, as these partner service activities are uncommon in Atlanta.

Experimental scenarios evaluated improvements to each partner services cascade step. Our first set of scenarios evaluated the impact of improving each step, alone and jointly. Counterfactual scenarios outcomes were compared to a “Missing = No” base scenario (**Table 1**) that calculated CDC partner services metrics assuming missing data represented a failure. Sensitivity analyses explored the alternative “complete case” assumption that excluded missing data (potentially overestimating success). Second, to understand the optimal impact of partner services, we simulated scenarios with all metrics at their maximum values. Third, we estimated the impact of more realistic improvements for the upstream steps, in which 90% of index patients and 25% or 50% of their partners were reached.

### Calibration, Simulation, and Analysis

We calibrated the model with a Bayesian approach that defined prior distributions for uncertain parameters and fit the model to surveillance data on diagnosed HIV, NG, and CT incidence for the target population (**Appendix S13**). After calibration, we simulated the model 500 times for each scenario and summarized the distribution of results with medians and 95% simulation intervals (SI). The time horizon for all scenarios was 10 years.

Our primary outcomes were HIV incidence per 100 person-years at risk (PYAR) in the last year of the time horizon and cumulative incidence across the horizon. We calculated the number and percent of infections averted (NIA and PIA, respectively) relative to the cumulative incidence in the base scenario. To estimate the efficiency of some experimental scenarios, we calculate the number needed to treat (NNT) as the number of index patients needing to start partner services to avert one new HIV infection. To understand the HIV prevention mechanisms, we calculated cross-sectional HIV care, prevention, and partner services cascade metrics.

## RESULTS

**Table 2** summarizes our base and primary experimental scenarios. In the base scenario, the point HIV incidence among Atlanta MSM was 1.16 per 100 PYAR (95% SI: 0.91, 1.44) at year 10, with 1,090 cumulative infections over 10 years. Improvements to each individual partner services step, while holding the values of other steps constant, had negligible impact on HIV incidence, with less than 1% of infections averted even when steps were individually set to their optimal (maximum) value.

**Table 2.**
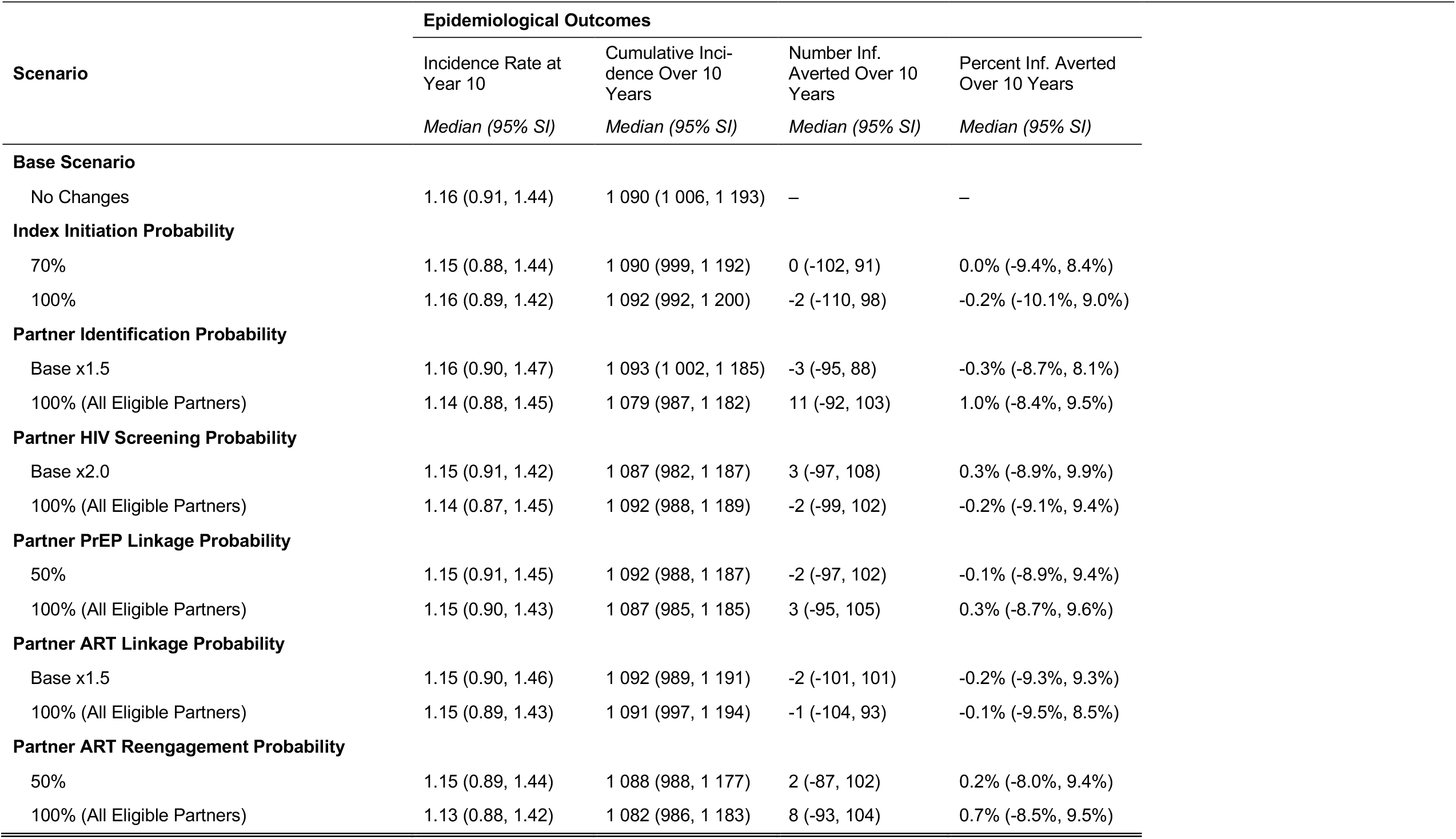
Impact of Changes to Individual Partner Services Cascade Step Changes on Point and Cumulative HIV Incidence

| Scenario | Epidemiological Outcomes |  |  |  |
| --- | --- | --- | --- | --- |
|  | Incidence Rate at Year 10 | Cumulative Incidence Over 10 Years | Number Inf. Averted Over 10 Years | Percent Inf. Averted Over 10 Years |
|  | *Median (95% SI)* | *Median (95% SI)* | *Median (95% SI)* | *Median (95% SI)* |
| **Base Scenario** |  |  |  |  |
| No Changes | 1.16 (0.91, 1.44) | 1 090 (1 006, 1 193) | – | – |
| **Index Initiation Probability** |  |  |  |  |
| 70% | 1.15 (0.88, 1.44) | 1 090 (999, 1 192) | 0 (-102, 91) | 0.0% (-9.4%, 8.4%) |
| 100% | 1.16 (0.89, 1.42) | 1 092 (992, 1 200) | -2 (-110, 98) | -0.2% (-10.1%, 9.0%) |
| **Partner Identification Probability** |  |  |  |  |
| Base x1.5 | 1.16 (0.90, 1.47) | 1 093 (1 002, 1 185) | -3 (-95, 88) | -0.3% (-8.7%, 8.1%) |
| 100% (All Eligible Partners) | 1.14 (0.88, 1.45) | 1 079 (987, 1 182) | 11 (-92, 103) | 1.0% (-8.4%, 9.5%) |
| **Partner HIV Screening Probability** |  |  |  |  |
| Base x2.0 | 1.15 (0.91, 1.42) | 1 087 (982, 1 187) | 3 (-97, 108) | 0.3% (-8.9%, 9.9%) |
| 100% (All Eligible Partners) | 1.14 (0.87, 1.45) | 1 092 (988, 1 189) | -2 (-99, 102) | -0.2% (-9.1%, 9.4%) |
| **Partner PrEP Linkage Probability** |  |  |  |  |
| 50% | 1.15 (0.91, 1.45) | 1 092 (988, 1 187) | -2 (-97, 102) | -0.1% (-8.9%, 9.4%) |
| 100% (All Eligible Partners) | 1.15 (0.90, 1.43) | 1 087 (985, 1 185) | 3 (-95, 105) | 0.3% (-8.7%, 9.6%) |
| **Partner ART Linkage Probability** |  |  |  |  |
| Base x1.5 | 1.15 (0.90, 1.46) | 1 092 (989, 1 191) | -2 (-101, 101) | -0.2% (-9.3%, 9.3%) |
| 100% (All Eligible Partners) | 1.15 (0.89, 1.43) | 1 091 (997, 1 194) | -1 (-104, 93) | -0.1% (-9.5%, 8.5%) |
| **Partner ART Reengagement Probability** |  |  |  |  |
| 50% | 1.15 (0.89, 1.44) | 1 088 (988, 1 177) | 2 (-87, 102) | 0.2% (-8.0%, 9.4%) |
| 100% (All Eligible Partners) | 1.13 (0.88, 1.42) | 1 082 (986, 1 183) | 8 (-93, 104) | 0.7% (-8.5%, 9.5%) |

**Supplemental Table 3** demonstrates why: individual step changes had limited impact on the HIV prevention and care cascade metrics of PrEP coverage and HIV VL suppression. **Supplemental Tables 4–5** subsequently demonstrate why *those* cascade metrics did not change. In the base scenario, over 10 years, the number of index patients reached was 622, who reported on 2,694 partners, of whom 273 were identified, of whom 92 HIV-screened, of whom 19 screened HIV-positive, of whom 6 were immediately linked to ART. It was only the MSM who reached this final step who had the potential to lower community VL, which could reduce the population transmission rate. When instead we assumed that all partners were identified, but everything else equal, ART linkage would increase to 63 partners; assuming optimal PrEP linkage, but everything else equal, 37 partners would start PrEP. These counts are too small to make a meaningful impact on PrEP coverage and VL suppression. **Supplemental Table 1** shows that limited impact was *not* due to the choice of calibration scenario, with no difference in incidence in the complete case scenario. In fact, when we simulated a scenario without partner services, there was effectively no difference in incidence, suggesting a minimal impact of partner services as currently delivered.

**Table 3** shows the projected impact under optimal conditions, in which both upstream partner services steps (index identification and partner identification) and downstream steps (PrEP, ART linkage, ART reengagement) were set to their maximum values. **Supplemental Figure 1** visualizes the HIV incidence time series across these scenarios. Here again, when changes were limited to upstream steps the impact was minimal (1.0% infections averted); however, as downstream services were added, the impact grew: 5.5% PIA when all partners were screened, 10.7% when all indicated partners were provided PrEP, 5.4% when newly diagnosed HIV-positive partners were linked to ART, 17.6% when previously diagnosed HIV-positive out of ART care were relinked to ART, and 21.7% when all of these downstream services were optimized. **Table 3** also provides the NNT, which ranges from 7.7 when upstream steps are optimized, down to 1.3 when all steps are optimized. The mechanisms for the HIV prevention impact are shown in **Supplemental Table 6**: PrEP coverage increased by 3 percentage points and HIV VL suppression increased by 6 percentage points. The mechanisms for these changes are demonstrated in **Supplemental Tables 7–8**: in our optimal scenario, the number of partners identified was 4,082, of whom 2,477 were HIV screened, of whom 1,454 started PrEP, 462 newly started ART, and 580 restarted ART.

**Table 3.** Impact of Changes to Partner Services on Point and Cumulative HIV Incidence, at Theoretical Maxima of Cascade Steps

| Scenario | Epidemiological Outcomes |  |  |  |  |
| --- | --- | --- | --- | --- | --- |
|  | Incidence Rate at Year 10 | Cumulative Incidence Over 10 Years | Number Inf. Averted Over 10 Years | Percent Inf. Averted Over 10 Years | NNT (Number of Indexes Started) |
|  | *Median (95% SI)* | *Median (95% SI)* | *Median (95% SI)* | *Median (95% SI)* | *Median (95% SI)* |
| **Base Scenario** |  |  |  |  |  |
| No Changes | 1.16 (0.91, 1.44) | 1 090 (1 006, 1 193) | – | – | – |
| **Counterfactual Scenarios** |  |  |  |  |  |
| All Index and All Partners Identified | 1.14 (0.90, 1.45) | 1 079 (983, 1 179) | 11 (-89, 107) | 1.0% (-8.2%, 9.8%) | 7.71 (-182.00, 362.05) |
| + All Partners HIV Screened | 1.10 (0.83, 1.37) | 1 030 (941, 1 123) | 60 (-33, 149) | 5.5% (-3.0%, 13.7%) | 5.93 (-53.91, 100.25) |
| + All Eligible Partners Provided PrEP | 1.02 (0.79, 1.26) | 973 (898, 1 058) | 117 (32, 192) | 10.7% (2.9%, 17.6%) | 3.38 (1.61, 12.77) |
| + All Eligible Partners Linked to ART | 1.08 (0.86, 1.38) | 1 031 (941, 1 131) | 59 (-41, 149) | 5.4% (-3.8%, 13.7%) | 6.15 (-76.53, 73.07) |
| + All Eligible Partners Relinked to ART | 0.91 (0.70, 1.15) | 898 (822, 973) | 192 (117, 268) | 17.6% (10.7%, 24.6%) | 1.81 (1.01, 3.63) |
| + All Eligible Linked and Relinked to ART | 0.90 (0.69, 1.16) | 894 (822, 972) | 196 (118, 268) | 18.0% (10.8%, 24.6%) | 1.76 (1.04, 3.45) |
| + All Partner Services at Maxima | 0.87 (0.65, 1.10) | 853 (778, 928) | 237 (162, 312) | 21.7% (14.9%, 28.6%) | 1.32 (0.78, 2.35) |

**Figure 1** visualizes the general relationship between the improvement in the two upstream steps (index and partner initiation probabilities) at current versus improved levels of downstream step service provision. The left panel shows that there is no impact on infections averted when scaling up one or both upstream identification steps without improvements in downstream steps. The right panel shows that when downstream services are first optimized, improving upstream steps yields a general linear increase in infections averted after some base provision of each. In summary, upstream steps were not impactful without improvement in downstream service provision, and downstream steps were maximized only with improvements to upstream steps.

**Figure 1.**
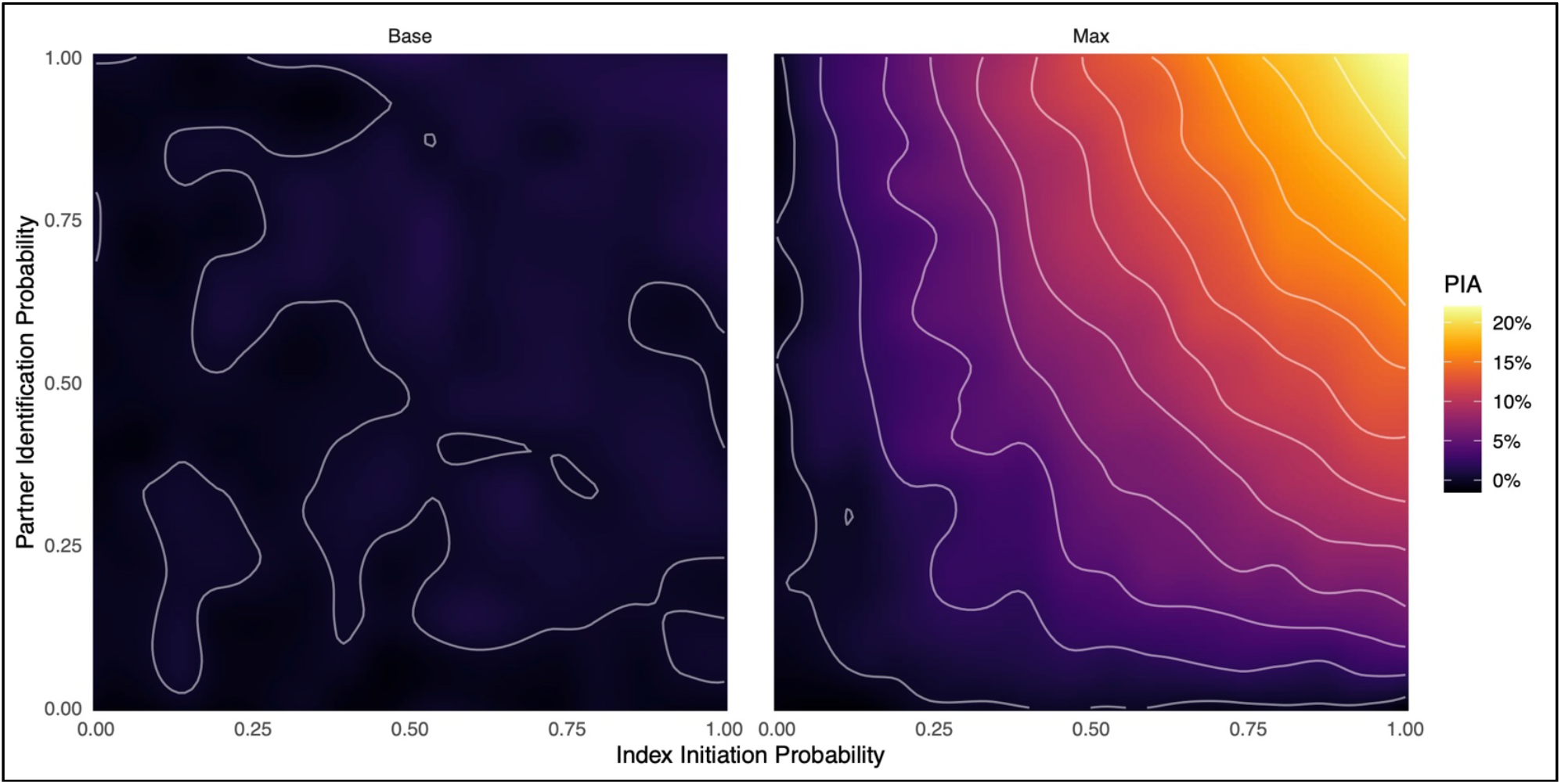
General relationship between improved partner services index initiation and partner identification probabilities on the 10-year percent of infections averted (PIA) compared to the base (calibrated) model scenario. Panel A (left) demonstrates the relationship under base downstream services (HIV screening, PrEP, and ART), whereas Panel B (right) shows the relationship when all downstream services are set to their theoretical maxima. Improvements to index or partner engagement have no discernable impact on the PIA under base downstream services but are associated with upwards of 20% infections averted when downstream services are maximized.

**Figure 2** and **Supplemental Tables 9–15** estimate the impact of partner services under more realistic scenarios in which 90% of index patients were identified and either 25% or 50% of partners were identified. While the impact on HIV incidence under these scenarios was not as high as the optimal scenarios, notable HIV prevention benefits (2–8% of infections averted) were achieved. **Figure 2** shows the relationship between improvements in HIV screening and improvements in either PrEP linkage or ART reengagement. Like **Table 3**, there was an overall larger relative benefit of ART service engagement than PrEP engagement. This was further demonstrated in the scenarios in **Supplemental Figure 2**, in which we observed greater synergy between HIV screening and ART scale-up than PrEP scale-up.

**Figure 2.**
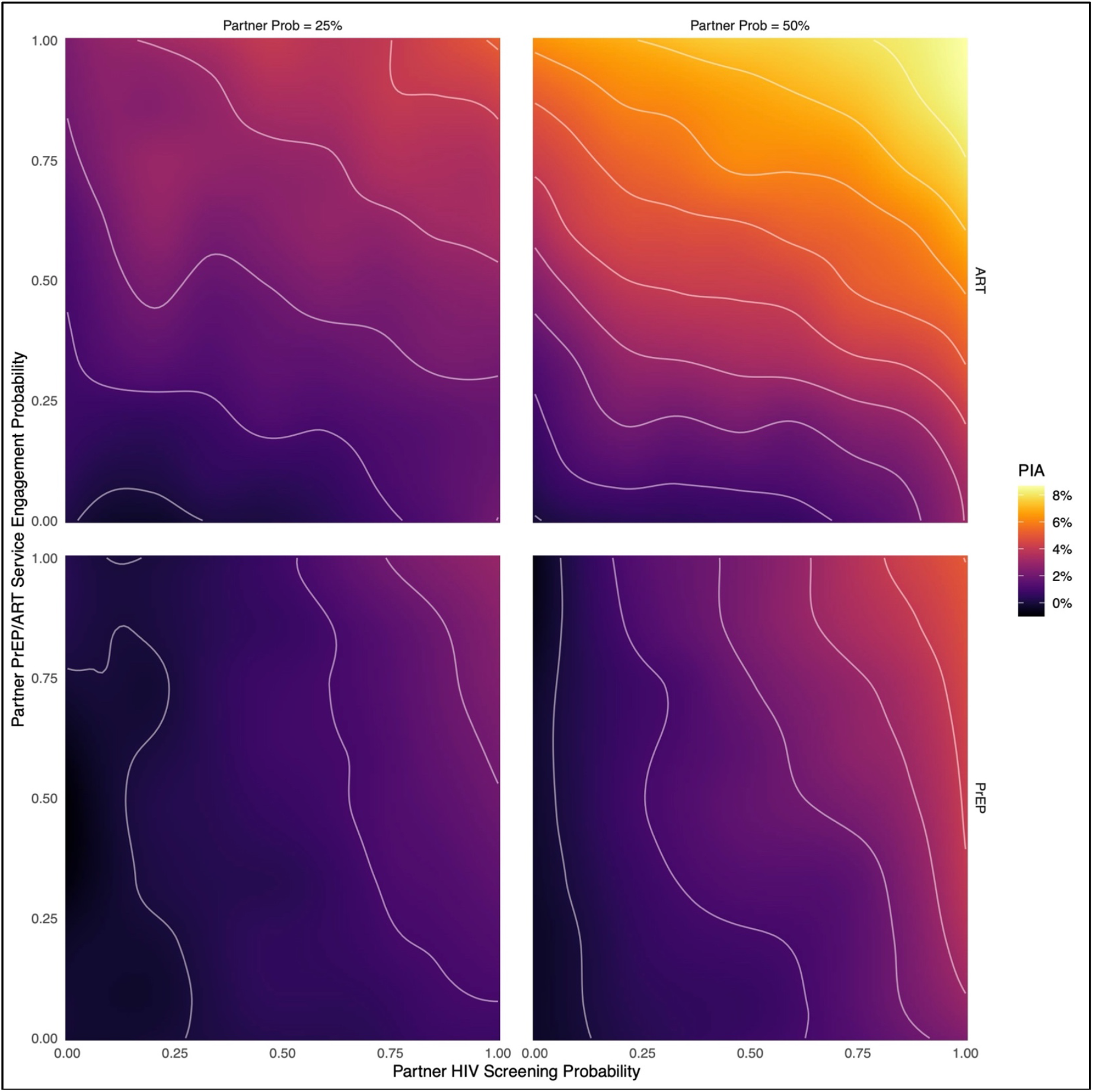
General relationship between improved partner HIV screening and either partner ART linkage/reengagement for HIVinfected partners (top panels) or PrEP initiation for HIV-negative partners (bottom panels) on the 10-year percent of infections averted (PIA) compared to the base (calibrated) model scenario, assuming 90% of index patients are identified and either 25% of their partners (left panels) or 50% of their partners (right panels) are identified. Higher levels of HIV screening are associated with greater HIV prevention, but ART engagement had a greater relative effect than PrEP initiation. The total benefit was greater when the partner identification was higher due to a larger number of identified partners who eligible for ART or PrEP.

## DISCUSSION

In this study, we estimated the population impact of HIV partner services in the context of our modern HIV biomedical prevention era of increased availability of highly effective antiretroviral HIV prevention tools. We found that HIV partner services, as currently delivered in our target population, provided clear clinical benefits to newly diagnosed index patients and their sexual partners. However, the intervention had limited impact on HIV transmission due to the steep drop from upstream cascade steps (identifying index patients and their partners) to delivery of downstream transmission-disrupting services (ART and PrEP). Improvements only to upstream steps had a negligible impact because of the limited delivery of downstream services. To achieve population prevention benefit, therefore, substantial but achievable improvements in the upstream steps of partner services must be paired with similar improvements in downstream service scale-up.

The success of partner services has traditionally been measured in terms of process metrics of partners identified and screened.^5,23^ However, it is unclear how these metrics translate into HIV incidence reduction. Mathematical modeling can help bridge the gap to estimate these quantities. However, models require realistic representations of sexual networks and the partner identification and HIV screening processes across those networks. Prior partner services models have used more simplistic representations that represent partner services as general increases in HIV diagnosis rates.^24^ This could overestimate the impact of partner services by failing to track partnership dyads that inherently limit the reach of this program. Our model builds upon the robust network structure representation that avoids this problem.^14^

Our study highlights the important contribution that partner services could make towards HIV prevention, with an estimated 22% of HIV infections averted under optimal service levels. Achieving these gains, however, will require substantial further investments to increase the size and speed of HIV partner services. This program have been weakened by the COVID-19 pandemic, which reallocated many disease intervention specialists (DIS) who conduct this work away from HIV and towards COVID contract tracing.^25^ In promising recent developments, after years of flat or declining funding for the public health workforce assigned to partner services, the U.S. American Recue Plan Act of 2021 has included $1 billion to expand and enhance DIS staff for HIV/STI partner services over the next five years.^26^ With this, some of the optimistic targets in our study may become feasible.

Our study also demonstrates the excellent efficiency (i.e., low NNT) of partner services, which translates into lower resources per infection averted. For example, we estimated an NNT of 1.3 index patients must be engaged to prevent one HIV infection under optimal delivery. This high efficiency for partner services was achieved by targeting HIV prevention services directly to areas of the sexual network with the highest HIV transmission potential. In comparison, we have estimated the NNT for individually targeted HIV PrEP in the same target population to be 20–30.^27^ A network-based cost-effectiveness analysis is suggested to compare investments in these two interventions as their costs and durability also differ.

The primary mechanism for partner services to impact HIV incidence is by reducing the time at each stage along the sequence from infection to diagnosis to ART initiation to VL suppression for sexual partners. The first step, HIV diagnosis, has been demonstrated to be a bottleneck in this sequence,^6^ delaying HIV VL suppression by years. Targeted HIV screening within partner services does fill a gap in individual-focused routine HIV screening efforts,^28^ but that does not necessarily translate to population prevention benefits. In our model, the accelerated HIV screening achieved through partner services by itself did not have a major impact, both because the current undiagnosed fraction was relatively low and the numbers initiating ART after testing through partner services were small. Still, if HIV screening for partners were maximized, our model suggests a prevention benefit of up to 5.5% of infections.

Efforts to further integrate delivery of ART and PrEP within partner services would also improve the overall impact of this program. Health jurisdictions like Seattle have used partner services to identify and link partners to HIV PrEP during time periods that are, by definition of being a contact with a HIV-infected person, high risk.^4^ A network intervention in Chicago successfully used partner services to link high-risk young persons to PrEP care.^29^ For ART, we found that partner services efforts to identify partners who are known to be HIV-positive but who have fallen out of HIV care could have a strong impact, consistent with prior research.^30^ Our study suggests that prioritizing ART reengagement for partners self-reported as HIV-positive but not in care had the overall greatest population benefit in HIV prevention (18% of infections averted).

## Limitations

First, we represented HIV partner services as a single type that would be provided by disease intervention specialists through local health authorities. In reality, alternative versions of partner services, both formal and informal, are provided by clinicians diagnosing index patients and even index patients themselves.^1^ However, the empirical data to differentiate these versions is too sparse to support a model. Including these alternative versions could expand the potential reach of partner services, but likely not to a degree that would substantially alter our conclusions.^31^ Second, we assumed that partner services only influenced the population force of infection through modifications to PrEP and ART coverage. Partner services may also have the ability to reduce HIV acquisition or transmission rates among identified partners through behavioral counseling (e.g., to increase condom use)^2^ or through linkage to testing and treatment for bacterial STIs.^32^ However, the causal evidence for these other prevention mechanisms are weak. Finally, although we represented the co-circulation of NG and CT in our model, we do not currently use an incident STI diagnosis as a trigger for HIV partner services. That was outside the current scope of this study, although is planned for future analyses. Including STIs to identify index patients could increase the overall index patient pool that could subsequently be elicited for partners who could benefit from HIV prevention services.^33^

## Conclusions

HIV partner services in the United States is at a crossroads. The new *Ending the Epidemic* plan^34^ calls for meeting ambitious HIV prevention goals in the next decade through a combination of public health efforts and intervention approaches. In the biomedical HIV prevention era, however, traditional partner services is complimented by a variety of individual-, partner-, network-, and community-based approaches to increase the coverage of antiretroviral medication to both persons with HIV (ART) and those without HIV at ongoing risk for HIV transmission or acquisition (PrEP).^35^ Our current study demonstrates the optimal reach of HIV partner services in this context, but also estimates its currently limited role in population health without substantial investments in the size and scope of this program. Recent increases in resources for partner services^26^ may help overcome these limitations. Our study can help guide the optimization of partner services to maximize its impact.

## Data Availability

All data produced in the present study are available upon reasonable request to the authors.

